# Diagnostic Performance and Regulatory Readiness of Dengue Rapid Diagnostic Tests Commercially Available in the United States

**DOI:** 10.1101/2025.07.23.25332048

**Authors:** Hans Desale, Forrest K Jones, Frances Vila-Hereter, Jessica Carrion, Manuela Beltran, Candimar Colon-Sanchez, Cindia Diaz, Genesis Cintron-De Leon, Camille Rivera Avilés, Maria Burgos-Garay, Carolynn DeByle, Roxana M. Rodríguez Stewart, Angel J. Rivera, Eliezer Rovira-Diaz, Laura Adams, Gabriela Paz-Bailey, Gilberto A. Santiago, Freddy A. Medina

## Abstract

Dengue is an emerging public health threat in the United States, and currently, no FDA-approved rapid diagnostic test (RDT) is available. A reliable RDT could support timely clinical management and outbreak response. We evaluated nine commercially available dengue RDTs using 197 stored serum samples: 99 from acute laboratory-confirmed dengue cases, 88 negative by RT-PCR, IgM ELISA, and NS1 ELISA, and 10 from acute confirmed Zika cases. Test performance varied, with sensitivity ranging from 36%–82% and specificity from 78%–100%. The CTK Biotech and Artron tests showed the highest sensitivity (80% and 82%, respectively), but only CTK Biotech maintained high specificity (98%). Cross-reactivity with Zika virus was low (0%–20%). These findings suggest that RDTs could enhance dengue detection in clinical settings, though further validation under field conditions is warranted, and some of these tests may be suitable for regulatory approval, which could lead to broader availability and use.

## Introduction

Dengue is a mosquito-borne disease of global health concern, caused by any of the four closely-related dengue viruses (DENV): DENV-1, DENV-2, DENV-3, DENV-4. In 2024, the World Health Organization reported the highest dengue case numbers on record, exceeding 14 million globally (*1*). Dengue remains a public health challenge in the United States, affecting people living in endemic areas as well as travelers to other countries with DENV transmission (*2*). In the United States, dengue transmission is considered frequent or continuous in six jurisdictions, including the U.S. territories of American Samoa, Puerto Rico, and the U.S. Virgin Islands, and the freely associated states of the Federated States of Micronesia, the Republic of Marshall Islands, and the Republic of Palau (*3*). In 2024, dengue outbreaks were declared in the U.S. Virgin Islands and Puerto Rico, with 6,291 confirmed and probable cases reported from Puerto Rico, 52% of which required hospitalization (*4*). Local transmission has also been reported in Florida, California, Texas, Hawaii, and Arizona (*5–8*). Additionally, hundreds of dengue cases among travelers in the U.S. are identified annually, with >3,500 travel associated cases reported in 2024 (*9*).

Dengue symptoms overlap with many other acute febrile illnesses, creating challenges for healthcare providers and potentially resulting in delays in initiating appropriate clinical management if other etiologies are suspected (*10*). Although clinicians are encouraged to start dengue clinical management without waiting for laboratory confirmation, accurate and timely diagnosis can be particularly informative in areas where dengue is less common. Early detection, followed by prompt clinical care with fluid replacement, can significantly reduce the risk of severe disease and mortality. Current acute dengue testing typically targets viral RNA, the secreted non-structural protein 1 (NS1), or anti-dengue IgM antibodies produced by the host immune response. During the first week of illness, NS1 antigen is typically detectable from symptom onset and may remain detectable for up to 9 days or longer, while IgM antibody begins to rise around day 3–5 and persists for weeks or months (*11*). While laboratory-based diagnostic tests provide high accuracy, they require specialized equipment, trained personnel, and processing time, forcing providers to rely on clinical judgment for patient management decisions.

Rapid diagnostic tests (RDTs) offer a potential solution by enabling easy-to-use, point-of-care results. Currently, no commercially available dengue RDTs have regulatory clearance or approval for diagnostic testing in the United States. This study comprehensively assesses the diagnostic performance of dengue RDTs commercially available in the United States.

## Methods

We selected 200 stored serum samples from specimens collected from a hospital-based enhanced dengue surveillance system (*12*), passive arboviral surveillance (*13*), and a household-based serosurvey among asymptomatic individuals (*14, 15*), all of which occurred in Puerto Rico. Sample size was constrained by the number of serum samples with sufficient volume to test all manufacturer RDTs (n=200). We used a quota sampling approach to randomly select approximately 100 dengue-positive samples, aiming for at least 50 positive by RT-PCR and 50 positive by IgM ELISA. Among RT-PCR–positive samples, we aimed for an even distribution of serotypes, with a target of roughly 12 samples per serotype to ensure broad representation. Negative samples came from randomly selecting a sample that were RT-PCR or IgM negative on the same plates. Individual samples were inspected and free from hemolysis. Samples were collected from 2018 to 2024, except for DENV-4 samples, which were collected in 2014 due to the absence of DENV-4 circulation during that period. Sample integrity was preserved through controlled freeze-thaw procedures, and reference tests were repeated on samples to confirm stability.

Reference testing was conducted using three FDA-approved dengue diagnostic tests: the CDC DENV-1-4 Real-time rRT-PCR Multiplex Assay (*16*), InBios DENV Detect™ NS1 ELISA (*17*), and InBios DENV Detect™ IgM Capture ELISA (*17*), following manufacturer recommended protocols. Among the 200 samples, three samples were ultimately not included because IgM ELISA results were inconsistent upon repeat testing, suggesting that sample stability was not maintained for those samples. The final sample set included 99 DENV-positive, 88 DENV- and ZIKV-negative, and 10 ZIKV-positive samples. RDT results were interpreted according to the antigen and antibody targets of each test: a positive result was defined as detection of NS1 antigen and/or IgM antibody in combination RDTs, or detection of NS1 alone in NS1-only RDTs. A negative result was defined as the absence of both NS1 and IgM in combination RDTs, or absence of NS1 in NS1-only RDTs. This definition reflects the closest approximation to true dengue infection status and facilitates estimation of each RDT’s sensitivity and specificity in identifying true dengue cases. To measure potential cross-reactivity with ZIKV, reference testing was performed using the CDC ZIKV IgM ELISA (*18*) and the CDC Trioplex Real-time RT-PCR Assay (*19*).

A global market analysis identified 59 manufacturers offering commercially available dengue rapid tests. This list was reduced to 9 manufacturers who sold their products in the United States and were available for the evaluation (Supplemental Figure 1). Of the commercially available dengue RDTs evaluated, three were designed for dengue NS1 antigen detection only: Abbexa Dengue Virus NS1 Antigen Rapid Test Kit (Abbexa), Biopanda Dengue NS1 Rapid Test Cassette (Biopanda), and InBios Dengue NS1 Detect™ Rapid Test (InBios). The remaining six were combination tests for dengue NS1 antigen and IgM antibody detection: Artron Dengue IgG/IgM & NS1 Test (Artron), Diagnostic Automation/Cortez Diagnostics OneStep Duo Panel (Cortez Diagnostics), Creative Diagnostics Combo Rapid Test (Creative Diagnostics), CTK OnSite Duo Dengue Ag-IgG/IgM Rapid Test (CTK Biotech), LumiQuick Diagnostics Duo Panel (LumiQuick Diagnostics), and MP Diagnostics MULTISURE Dengue Ab/Ag Rapid Test (MP Diagnostics) (Supplemental table 1). IgG results were excluded from analysis, as they represent past infection and do not contribute to acute dengue diagnosis.

All RDTs were performed and interpreted according to the manufacturer recommended instructions. Two independent readers interpreted each test result and discrepant results were resolved by a third reader. Testers and readers were blinded to both infection status and each other’s records. High-resolution photographs of all test results were taken to document findings. Invalid or indeterminate test results were excluded from the final analysis. This study was conducted and reported in accordance with the STARD (Standards for Reporting Diagnostic Accuracy Studies) guidelines (*20*).

Sensitivity was calculated as the proportion of reference-positive samples that tested positive by RDT. Specificity was calculated as the proportion of reference-negative samples that tested negative by RDT. Positive predictive values (PPV) and negative predictive values (NPV) were calculated using the estimates of sensitivity and specificity with different levels of infection prevalence. Percent agreement was calculated using the same formulas as sensitivity and specificity but was used to measure concordance between two diagnostic tests without designating a reference standard. To evaluate the impact of day post-onset distribution on test performance, adjusted sensitivity and specificity estimates were modeled by reweighting based on either a uniform DPO distribution or the DPO distribution observed in a hospital-based enhanced dengue surveillance system in Puerto Rico. Data analysis was performed using R version 4.4.0 (*21*) and figures produced using GraphPad Prism version 10.1.2 (*22*).

## Results

### Sample Characteristics

The study included 99 DENV-positive, 88 DENV-negative, and 10 ZIKV–positive serum samples selected to represent a range of diagnostic profiles and timepoints post-symptom onset (Table 1). Among the DENV-positive samples, 6 were positive by DENV IgM ELISA only, 14 by both DENV RT-PCR and DENV IgM ELISA, 29 by DENV RT-PCR and DENV NS1 ELISA, 10 by DENV RT-PCR alone, and 40 by all three reference assays. All DENV-negative and ZIKV-positive samples were negative by DENV RT-PCR, DENV NS1 ELISA, and DENV IgM ELISA.

**Table 1.** Demographic and diagnostic characteristics of serum samples by infection status from 2014–2024, Puerto Rico.

| Characteristic | No. (%) |  |  |
| --- | --- | --- | --- |
|  | DENV-positive, n = 99 | Negative, n = 88 | ZIKV-positive, n = 10 |
| DENV reference test results |  |  |  |
| InBios DENV Detect™ IgM Capture ELIS only (IgM ELISA) | 6 (6.1%) | 0 (0%) | 0 (0%) |
| CDC DENV-1-4 Real-Time rRT-PCR Multiplex Assay (DENV RT-PCR) and IgM ELISA | 14 (14.1%) | 0 (0%) | 0 (0%) |
| DENV RT-PCR and InBios DENV Detect™ NS1 ELISA (NS1 ELISA) | 29 (29.3%) | 0 (0%) | 0 (0%) |
| DENV RT-PCR only | 10 (10.1%) | 0 (0%) | 0 (0%) |
| DENV RT-PCR, IgM ELISA, and NS1 ELISA | 40 (40.4%) | 0 (0%) | 0 (0%) |
| Negative by DENV RT-PCR, NS1 ELISA, and IgM ELISA | 0 (0.0%) | 88 (100%) | 10 (100%) |
| Median age of individual at collection* | 24 (15, 37) | 35 (20, 47) | 41 (19, 48) |
| Distribution of DENV serotype |  |  |  |
| DENV-1 | 38 (38.4%) | – | – |
| DENV-2 | 12 (12.1%) | – | – |
| DENV-3 | 28 (28.3%) | – | – |
| DENV-4 | 15 (15.2%) | – | – |
| Unknown | 6 (6.1%) | – | – |
| Days between symptom onset and sample collection |  |  |  |
| 0 | 2 (2.0%) | 3 (3.4%) | 0 (0%) |
| 1 | 6 (6.1%) | 12 (13.6%) | 0 (0%) |
| 2 | 12 (12.1%) | 9 (10.2%) | 0 (0%) |
| 3 | 7 (7.1%) | 3 (3.4%) | 0 (0%) |
| 4 | 23 (23.2%) | 4 (4.5%) | 10 (100%) |
| 5 | 27 (27.3%) | 3 (3.4%) | 0 (0%) |
| 6 | 12 (12.1%) | 3 (3.4%) | 0 (0%) |
| 7 | 8 (8.1%) | 4 (4.5%) | 0 (0%) |
| 8+ | 2 (2.0%) | 7 (8.0%) | 0 (0%) |
| Unknown | 0 (0.0%) | 40 (45.5%) | 0 |
| Sample sources by surveillance project |  |  |  |
| Hospital-based enhanced surveillance | 75 (76.0%) | 52 (59.0%) | 1 (10.0%) |
| Passive dengue surveillance systems | 24 (24.0%) | – | 9 (90.0%) |
| Serosurvey among asymptomatic individuals | – | 36 (41.0%) | – |

\*Median (IQR).

The median patient age at the time of sample collection was 24 years (IQR: 15– 37) among DENV-positive participants, 35 years (IQR: 20–47) among DENV-negative participants, and 41 years (IQR: 22–48) among ZIKV-positive participants. All four DENV serotypes were represented in the DENV-positive group: DENV-1 (38.4%), DENV-2 (12.1%), DENV-3 (28.3%), and DENV-4 (15.2%). Serotype was unknown among the six samples that were only positive by DENV IgM ELISA. The highest proportion of DENV-positive samples were collected 4 or 5 days post-symptom onset (23.2% and 27.3% respectively). DENV-negative samples were more commonly collected earlier, with 13.6% collected on day 1 and 10.2% on day 2. All ZIKV-positive samples were collected on day 4 post-symptom onset. DENV-positive samples were obtained from symptomatic individuals either through hospital-based enhanced surveillance (76%) or passive dengue surveillance systems (24%). DENV-negative samples were collected from both hospital-based enhanced surveillance (59%), and serosurveys among asymptomatic individuals (41%). ZIKV-positive samples were primarily obtained through passive surveillance (90%), with the remaining 10% collected via enhanced surveillance.

### RDT Sensitivity and Specificity

The sensitivity and specificity of each RDT, calculated against the composite reference standard, varied widely (Figure 1, Table 2). Sample flow and index tests outcomes are provided using STARD flow diagrams (Supplemental Figures S3-11). CTK Biotech and Artron demonstrated the highest sensitivities at 80% (95% CI: 71–87%) and 82% (95% CI: 73–89%), respectively. CTK Biotech also exhibited a high specificity of 98% (95% CI: 92–100%), while Artron exhibited a lower specificity of 78% (95% CI: 68– 86). LumiQuick Diagnostics, Cortez Diagnostics, Biopanda, and InBios showed moderate sensitivity (ranging from 56% to 62%) with high specificity (92%–97%). Creative Diagnostics, MP Diagnostics, and Abbexa demonstrated lower sensitivities, ranging from 36% to 49%, although their specificities were among the highest (99%– 100%). To assess the added value of combination RDTs, we compared the diagnostic performance of individual NS1 and IgM components to their combined results (Table 2). Across most tests, sensitivity improved when components were combined, with the greatest increases observed for Artron (NS1: 74%, IgM: 53%, combined: 82%) and CTK Biotech (NS1: 58%, IgM: 52%, combined: 80%); whereas Creative Diagnostics and MP Diagnostics showed minimal gains from combining components. Specificity was typically highest for NS1 alone and declined slightly when IgM was added; for example, Artron dropped from 81% (NS1) and 95% (IgM) to 78% combined. However, some tests, including Creative Diagnostics, Cortez, and MP Diagnostics, maintained high specificity (>97%) across all formats.

**Figure 1.**
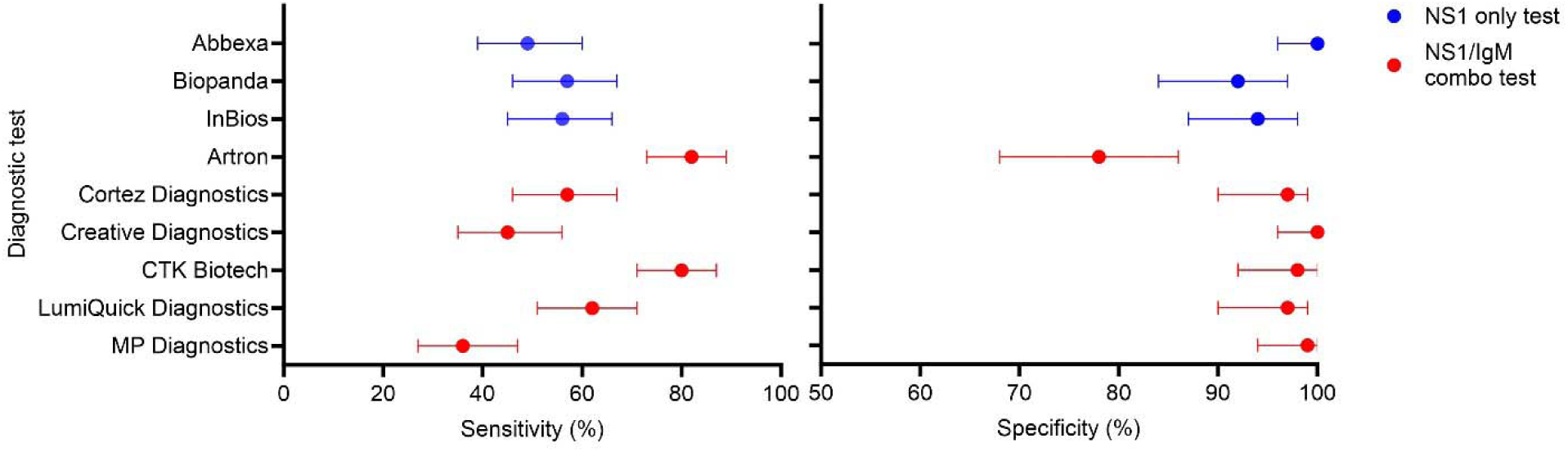
Sensitivity and specificity of dengue rapid diagnostic tests (RDTs) compared with a composite reference standard, with 95% confidence intervals. The composite reference standard defined dengue-positive samples as those positive by any FDA-approved RT-PCR, NS1, or IgM ELISA dengue test. Red dots indicate NS1/IgM combination RDTs; blue dots indicate NS1-only RDTs. Note different x-axis limits between graphs.

**Table 2.** Sensitivity and specificity of dengue rapid diagnostic tests compared to a composite reference standard.

| % (95% Confidence interval) |  |  |  |  |  |  |
| --- | --- | --- | --- | --- | --- | --- |
| Rapid diagnostic test | Sensitivity, n=99 |  |  | Specificity, n=88 |  |  |
|  | NS1 component only | IgM component only | NS1 & IgM combined components | NS1 component only | IgM component only | NS1 & IgM components combined |
| Abbexa* | 49% (39-60) | – | – | 100% (96-100) | – | – |
| Biopanda | 57% (46-67) | – | – | 92% (84-97) | – | – |
| InBios | 56% (45-66) | – | – | 94% (87-98) | – | – |
| Artron | 74% (64-82) | 53% (42-63) | 82% (73-89) | 81% (71-88) | 95% (89-99) | 78% (68-86) |
| Cortez Diagnostics | 42% (33-53) | 35% (26-46) | 57% (46-67) | 100% (96-100) | 97% (90-99) | 97% (90-99) |
| Creative Diagnostics | 43% (33-54) | 15% (9-24) | 45% (35-56) | 100% (96-100) | 100% (96-100) | 100% (96-100) |
| CTK Biotech | 58% (47-67) | 52% (41-62) | 80% (71-87) | 100% (96-100) | 98% (92-100) | 98% (92-100) |
| LumiQuick Diagnostics† | 48% (38-59) | 38% (28-48) | 62% (51-71) | 100% (96-100) | 97% (90-99) | 97% (90-99) |
| MP Diagnostics | 12% (6-20) | 25% (17-35) | 36% (27-47) | 100% (96-100) | 99% (94-100) | 99% (94-100) |

\* One Abbexa NS1 result was invalid and excluded; NS1 analyses are based on n = 98, while IgM analyses are based on n = 99..
† One LumiQuick Diagnostics IgM result was invalid and excluded; IgM analyses are based on n = 98, while NS1 analyses are based on n = 99..

Sensitivity was evaluated for each DENV serotype among DENV-positive samples with serotype available (n=93). Sensitivity varied across DENV serotypes, with notable differences by serotype (Figure 2, Supplemental Table 2). Overall, test performance was highest for DENV-1 and DENV-3, while sensitivities were generally lower for DENV-2 and DENV-4. CTK Biotech and Artron showed the highest sensitivities across all serotypes. LumiQuick Diagnostics and Cortez Diagnostics had moderate sensitivity, but performance was inconsistent across serotypes, with a marked drop in sensitivity for DENV-4. Creative Diagnostics, MP Diagnostics, and Abbexa exhibited the lowest sensitivities, particularly for DENV-2 and DENV-4.

**Figure 2.**
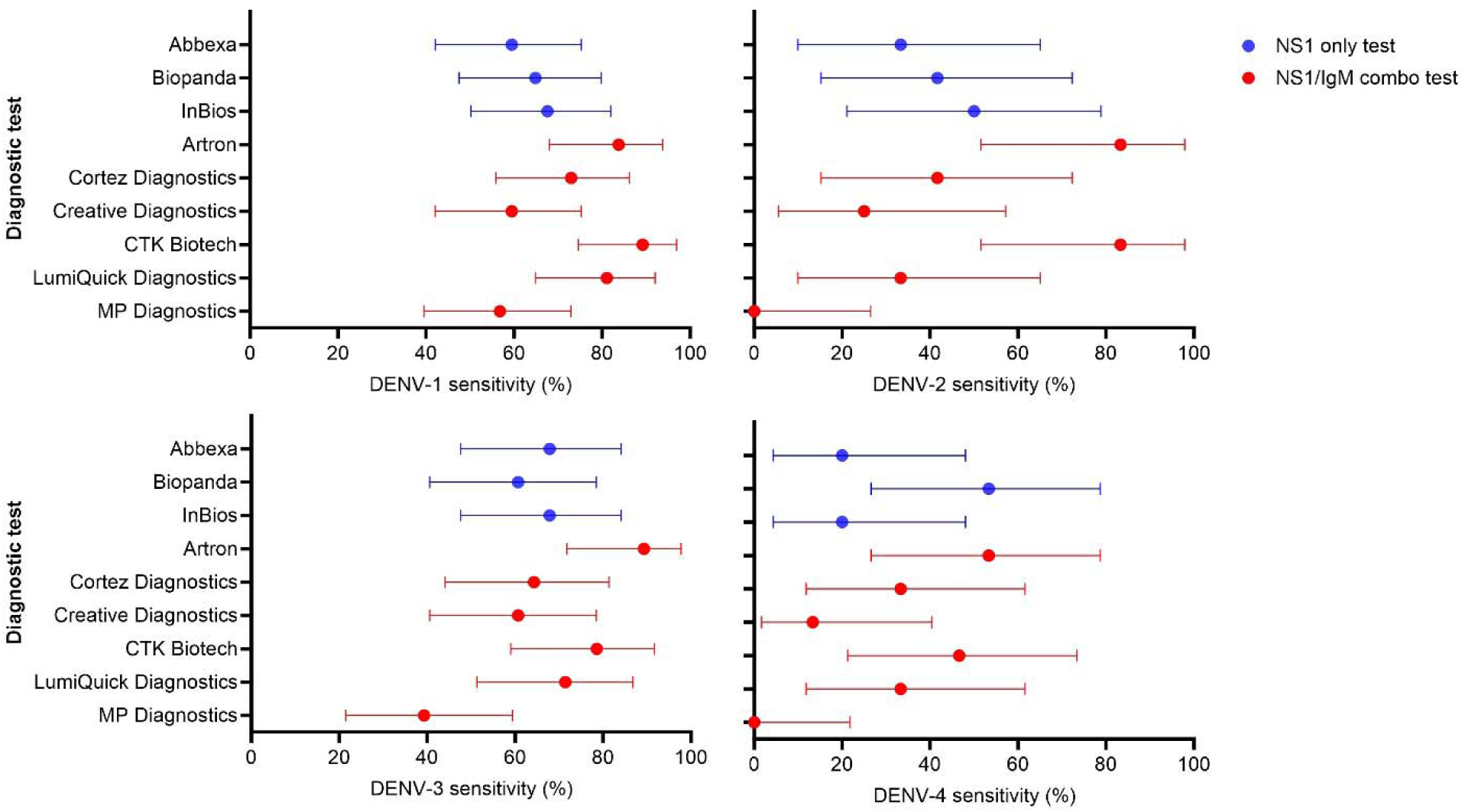
Sensitivity by serotype of dengue rapid diagnostic tests (RDTs) compared with a composite reference standard, with 95% confidence intervals. The composite reference standard defined dengue-positive samples as those positive by FDA-approved RT-PCR dengue test (n=93). Red dots indicate NS1/IgM combination RDTs; blue dots indicate NS1-only RDTs. DENV, dengue virus; DENV-1, DENV serotype 1; DENV-2, DENV serotype 2; DENV-3, DENV serotype 3; DENV-4, DENV serotype 4

Test sensitivity varied by day of specimen collection after symptom onset, with most RDTs demonstrating increased sensitivity at later time points (Figure 3). Across all tests, sensitivity was lowest within the first 1–2 days after symptom onset, gradually improving by days 4–6. CTK Biotech and Artron exhibited the highest overall sensitivity throughout the illness course. Biopanda showed strong early sensitivity but experienced a decline in later days post-onset, likely associated with waning NS1 levels. Cortez, Creative Diagnostics, and InBios showed moderate improvements in sensitivity over time after the first two days post symptom onset. Abbexa, LumiQuick Diagnostics, and MP Diagnostics had the lowest sensitivity in the early days post-onset but increased detection rates later in the illness course. After adjusting for time after symptom onset based on distributions observed in surveillance data, sensitivity estimates increased slightly for NS1-based test components, and decreased slightly for IgM-based test components; however, the relative performance of RDTs remained consistent, suggesting that observed differences reflect true test characteristics rather than artifacts of sample distribution (Supplemental Figure 2).

**Figure 3.**
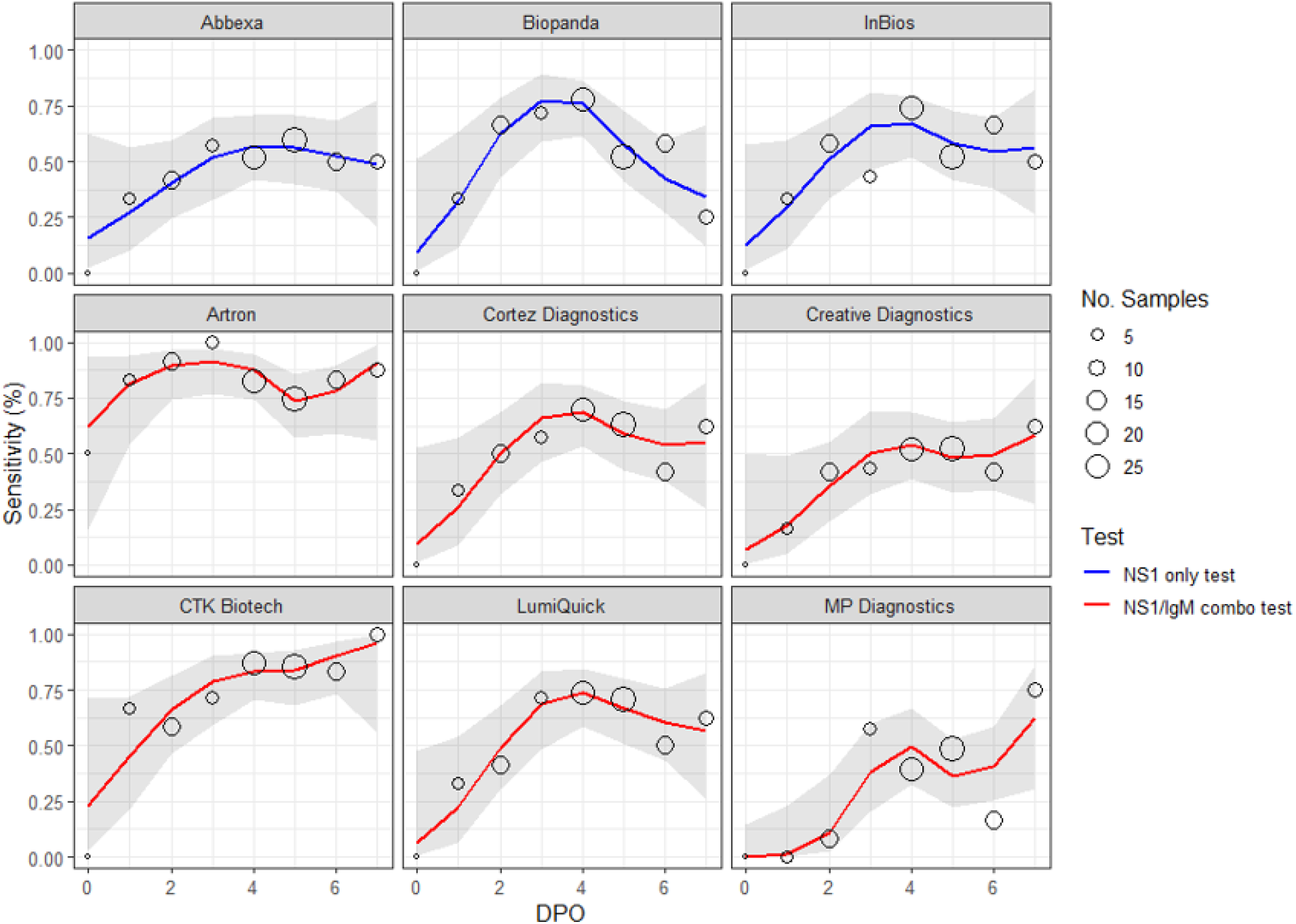
Time-varying sensitivity of dengue rapid diagnostic tests (RDTs) compared with a composite reference standard by day post-symptom onset (DPO). The composite reference standard defined dengue-positive samples as those positive by any FDA-approved RT-PCR, NS1, or IgM ELISA dengue test. Panels show sensitivity estimates modeled using a cubic spline for each RDT for DPO 0–7, with shaded 95% confidence intervals. Red lines indicate NS1/IgM combination RDTs; blue lines indicate NS1-only RDTs. Circle size indicates the sample size of tested samples at that DPO.

### RDT Predictive Values

Positive predictive values (PPV) and negative predictive values (NPV) were calculated for RDTs that met the threshold of at least 80% sensitivity and 95% specificity, which included CTK Biotech and Artron (Figure 4). PPV and NPV reflect a test’s practical utility, with PPV indicating the likelihood of a true positive and NPV the likelihood of a true negative based on the test result. Unlike sensitivity and specificity, which are inherent test properties, PPV and NPV are dependent on the prevalence of disease in the tested population. CTK Biotech consistently demonstrated higher PPV than Artron across all prevalence levels, whereas both tests maintained high NPV at lower dengue prevalence.

**Figure 4.**
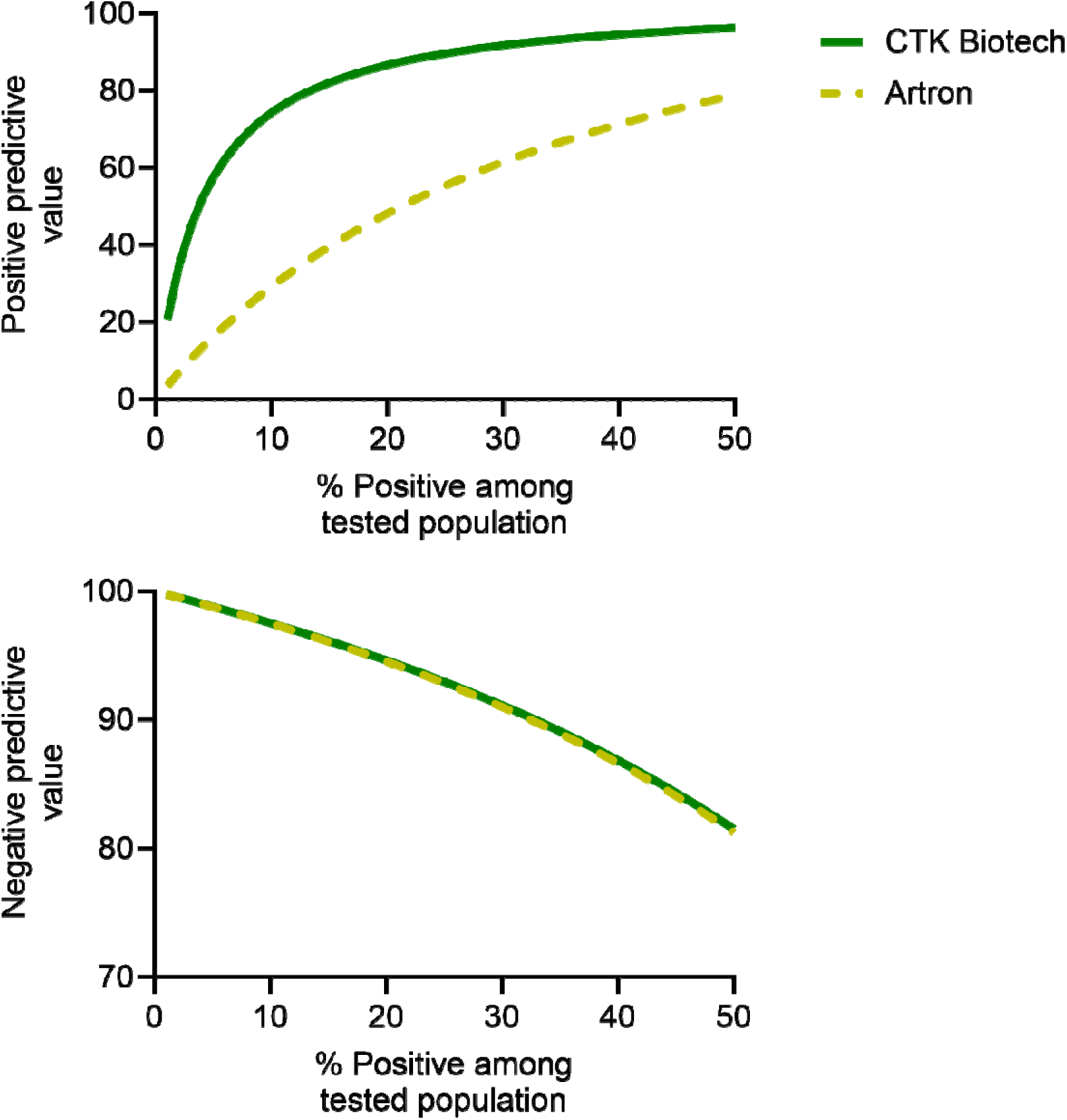
Predictive values for dengue rapid diagnostic tests (RDTs) with ≥80% sensitivity: Artron (gold dashes) and CTK Biotech (green line) compared with a composite reference standard, modeled as functions of the percentage of true dengue cases among those tested. The composite reference standard defined dengue-positive samples as those positive by any FDA-approved RT-PCR, NS1, or IgM ELISA dengue test. Note different y-axis limits between graphs.

### NS1/IgM Percent Agreement

In assessing positive percent agreement (PPA) for the NS1 RDT compared to the NS1 ELISA, Artron demonstrated the highest PPA at 91% (95% CI: 82–97%), closely followed by CTK Biotech at 81% (95% CI: 70–90%), while MP Diagnostics had the lowest at 17% (95% CI: 9–28%) (Table 3). Most RDTs showed high negative percent agreement (NPA) for NS1, with several tests—including CTK Biotech, LumiQuick Diagnostics, Cortez Diagnostics, Creative Diagnostics, MP Diagnostics, and Abbexa— reaching or exceeding 99%. For IgM, CTK Biotech had the highest PPA with the IgM ELISA at 80% (95% CI: 68–89%) and maintained high NPA at 98% (95% CI: 94–100%). Artron showed moderate IgM PPA at 70% and NPA at 90%. In contrast, Creative Diagnostics had the lowest IgM PPA at 23% (95% CI: 13–36%), despite 100% NPA.

**Table 3.** Percent agreement of dengue rapid diagnostic test components with respective NS1 and IgM ELISAs and cross-reactivity with Zika virus–positive samples.

| Manufacturer | % (95% Confidence interval) |  |  |  |  |  |
| --- | --- | --- | --- | --- | --- | --- |
|  | NS1 component-NS1<br>ELISA positive percent<br>agreement (n=69) | NS1 Component-NS1<br>ELISA negative percent<br>agreement (n=118) | NS1 Component<br>Zika Cross-Reactivity (n=10) | IgM Component-IgM<br>ELISA positive percent<br>agreement (n=60) | IgM Component-IgM<br>ELISA negative percent<br>agreement (n=127) | IgM Component<br>Zika Cross-Reactivity (n=10) |
| Abbexa* | 70% (57-80) | 99% (95-100) | 0% (0-31) | – | – | 0% (0-31) |
| Biopanda | 72% (60-83) | 89% (82-94) | 0% (0-31) | – | – | 0% (0-31) |
| InBios | 75% (64-85) | 93% (87-97) | 0% (0-31) | – | – | 0% (0-31) |
| Artron | 91% (82-97) | 77% (68-84) | 20% (3-56) | 70% (57-81) | 90% (83-95) | 0% (0-31) |
| Cortez<br>Diagnostics | 61% (48-72) | 100% (97-100) | 0% (0-31) | 47% (34-60) | 93% (87-97) | 10% (0-45) |
| Creative<br>Diagnostics | 62% (50-74) | 100% (97-100) | 0% (0-31) | 23% (13-36) | 100% (97-100) | 10% (0-45) |
| CTK Biotech | 81% (70-90) | 99% (95-100) | 0% (0-31) | 80% (68-89) | 98% (94-100) | 10% (0-45) |
| LumiQuick<br>Diagnostics† | 70% (57-80) | 100% (97-100) | 0% (0-31) | 56% (42-69) | 95% (89-98) | 10% (0-45) |
| MP Diagnostics | 17% (9-28) | 100% (97-100) | 0% (0-31) | 40% (28-53) | 99% (95-100) | 10% (0-45) |

\*One Abbexa NS1 result was invalid and excluded; positive percent agreement, negative percent agreement, and Zika cross-reactivity calculations are based on n=68, n=117, and n=9, respectively.
†One LumiQuick Diagnostics IgM result was invalid and excluded; positive percent agreement, negative percent agreement, and Zika cross-reactivity calculations are based on n=59, n=126, and n=9, respectively.

### Zika Cross-Reactivity

To evaluate cross-reactivity, each RDT was tested on 10 ZIKV samples. All samples tested positive in both the CDC ZIKV IgM ELISA and the CDC Trioplex Real-time RT-PCR Assay. A DENV-false positive was defined as a positive RDT result by NS1 and/or IgM in a ZIKV-positive sample. Most tests showed low levels of cross-reactivity, with false-positive rates ranging from 0% to 20% (Table 3). Artron had the highest observed cross-reactivity at 20% (95% CI: 3–56%), followed by CTK Biotech, LumiQuick Diagnostics, Cortez Diagnostics, Creative Diagnostics, and MP Diagnostics, each at 10% (95% CI: 0–45%). Abbexa, Biopanda, and InBios, all NS1-only RDTs, showed no false positives (0%, 95% CI: 0–31%). Among the tests with observed cross-reactivity, all false positives were due to the IgM component, except for Artron, which had both false positives in the NS1 component.

## Discussion

This evaluation of commercially available dengue RDTs revealed substantial variability in diagnostic performance, with CTK Biotech emerging as the only test demonstrating both high sensitivity (80%) and high specificity (98%), making it a promising candidate for clinical use. In addition to its diagnostic performance, CTK Biotech required a relatively low sample volume (65 µL), further supporting its feasibility in clinical settings. Several rapid diagnostic tests for COVID-19 and influenza have achieved FDA authorization with sensitivities ranging from 70%–90% and specificities above 90% (*23–25*), suggesting that the performance of CTK Biotech’s dengue RDT may fall within an acceptable range for regulatory consideration across infectious disease diagnostics.

This study demonstrated variability in the sensitivity and specificity of the NS1 and IgM components across different RDTs. Some tests, such as CTK Biotech, exhibited improved sensitivity with the addition of the IgM component while maintaining high specificity, suggesting a more balanced diagnostic performance. Others, like Artron, showed increased sensitivity due to the IgM component but at the expense of reduced specificity. Other assays, such as Creative Diagnostics, provided minimal additional diagnostic value from IgM. In clinical practice, optimal use of dengue RDTs depends not only on test performance but also on contextual factors such as timing of testing, local epidemiology, and co-circulating arboviruses. This evaluation found that RDT performance varied across the illness course; for example, CTK Biotech and Artron showed high overall sensitivity with more consistent performance across acute DPOs, while others had higher sensitivity only at certain timepoints, suggesting that test interpretation may depend on the day post-onset of testing, and selecting tests with consistent performance across DPOs may be preferable, especially when DPO is uncertain or imprecisely reported. Epidemiologic risk factors such as residence in or travel to endemic areas, or testing during outbreak settings, increase the pre-test probability of dengue and should inform interpretation of RDT results discordant with clinical suspicion. Predictive value modeling among the evaluated RDTs highlighted that modest reductions in specificity led to substantial declines in PPV, particularly in low-prevalence settings, while NPV remained high across all tests despite wide variability in sensitivity. Overall, the use of dengue RDTs in non-endemic areas among patients without travel or epidemiologic links is unlikely to be useful, given the low expected infection prevalence (<5%) and the high likelihood that positive results would be false positives. However, RDTs may be valuable in settings with known dengue virus transmission or among travelers returning from endemic areas with compatible symptoms. In the United States, RDT use should be guided by the clinical and epidemiologic likelihood of dengue, and confirmatory testing remains important when prevalence is low. The possibility of cross-reactive flaviviruses like Zika or West Nile Virus should always be considered when diagnosis dengue, especially when interpreting IgM results. Zika cross-reactivity was not frequent in this evaluation, and primarily limited to the IgM components of RDTs; only the Artron test showed cross-reactivity in its NS1 component. These findings highlight the need to interpret RDT results considering local disease circulation patterns and the specific antigen or antibody targets of each test. Integrating this information can improve diagnostic accuracy and guide appropriate follow-up testing when needed.

This study has several limitations. First, testing was performed on serum samples under laboratory conditions, whereas RDTs are intended for use with whole blood in clinical settings; future studies will focus on the performance of tests in their intended setting. Second, the small number of early post-onset samples constrained evaluation of RDT performance during the initial stages (days 0-1) of illness. This limitation arose because sample selection was intentionally designed to balance serotype distribution, day post-onset, and DENV-positive and -negative sample distribution to allow for precise estimation of both sensitivity and specificity. Samples were also selected to include enough NS1- and IgM-positive samples to ensure evaluation of combination RDTs. As a result, the observed performance may underestimate the true diagnostic potential of RDTs in the initial days post-symptom onset. Lastly, we did not investigate potential cross-reactivity with flaviviruses other than Zika virus.

In summary, this study provides a comprehensive evaluation of dengue RDTs commercially available in the United States, highlighting variability in diagnostic performance. While one test demonstrated both high sensitivity and specificity, others showed trade-offs between these measures. These findings suggest that certain RDTs may be suitable for supporting clinical decision-making among patients living in areas with local dengue transmission or with recent travel to dengue-endemic areas. As dengue incidence rises globally and local transmission expands in the United States, improving access to accurate point-of-care diagnostics approved by regulatory authorities remains a critical priority. Regulatory agencies are likely to require studies that include clinical samples from U.S.-based patients, which presents important challenges. This study underscores the difficulty of obtaining well-characterized samples across the full course of illness, by serotype, serostatus, and days post-onset— particularly for less common serotypes like DENV-4. These challenges will be even more pronounced in prospective studies conducted exclusively in the U.S. Allowing the inclusion of stored samples from international settings could significantly improve study feasibility and strengthen the evidence base for regulatory approval. Early coordination among manufacturers, public health programs, and regulatory agencies will be essential to ensure access to sufficient and appropriately diverse samples and to support robust study design.

## Data Availability

Raw data and code used in this study is available at: https://github.com/hdesale-2408/DENVRDTEvaluation_2024-2025.git

## Acknowledgements

We thank Dr. Joshua Wong for his clinical advice.

## Disclaimer

The findings and conclusions in this report are those of the authors and do not necessarily represent the views of the Centers for Disease Control and Prevention.

