## Supplemental Tables and Figures for "Diagnostic Performance and Regulatory Readiness of Dengue Rapid Diagnostic Tests Commercially Available in the United States"

Supplemental table 1. Characteristics of commercially available dengue rapid diagnostic tests evaluated in this study

| **Manufacturer** | **Product name** | **Catalog no.** | **Package insert, date** | **Analyte(s) detected** | **Recommended specimen type(s)** | **Time to result (minutes)** | **Required test volume (µL)** |
| --- | --- | --- | --- | --- | --- | --- | --- |
| Abbexa | Dengue Virus NS1 Antigen Rapid Test Kit | Abx090706 | Abbexa abx090706_ifu Version 1.0.1, Revision date 10-Nov-23 | NS1 Antigen | Whole blood, serum, plasma | 20 | 60 |
| Biopanda | Dengue NS1 Rapid Test Cassette | RAPG-DES-001 | Dengue NS1 Rapid Test Cassette RAPG-DES-001 Insert, Revision 5-12-2023 | NS1 Antigen | Whole blood, serum, plasma | 10-20 | 75 |
| InBios | Dengue NS1 *Detect*™ Rapid Test | DNS1-RD | InBios_900111-00-RUO-Dengue-NS1-Detect-Rapid-Test-Dry-printable-version Insert Part No 900111-00 | NS1 Antigen | Serum | 30-45 | 50 |
| Artron | Dengue IgG/IgM & NS1 Test | A03-24-322 | Arton_IFU_A03-24-322 Dengue Combo WB (cassette) Version 22-01, Revision Sep 2020 | NS1 Antigen, IgG Antibody, IgM Antibody | Whole blood, serum, plasma | 10-30 | 40 |
| Cortez Diagnostics | OneStep Dengue NS1 Antigen & IgG/IgM Antibody Duo Panel Rapicard™ InstaTest | 173112-25-23 | RUO-Dengue_NS1&IgG-IgM_Duo_RapiCard-173112-25-23 (03-01-2023), Revision 03-01-2023 | NS1 Antigen, IgG Antibody, IgM Antibody | Whole blood, serum, plasma | 20 | 125 |
| Creative Diagnostics | Dengue IgG/IgM/NS1 Combo Rapid Test Kit | DTS349-2402 | Creative Diagnostics DTS349-2402 Instructions for Use | NS1 Antigen, IgG Antibody, IgM Antibody | Whole blood, serum, plasma | 10-15 | 20 |
| CTK Biotech | OnSite Duo Dengue Ag-IgG/IgM Rapid Test-Cassette | R0062C | OnSite Duo Dengue Ag-IgG/IgM Rapid Test-Cassette Ref R0062C Instructions for Use, Copyright 2020 | NS1 Antigen, IgG Antibody, IgM Antibody | Whole blood, serum, plasma | 20-25 | 65 |
| LumiQuick Diagnostics | Dengue NS1 Antigen & IgG/IgM Antibody Duo Panel | 71087 | LumiQuick IgM-IgG-NS1 DCR 24-028, Revision 06-27-2024 | NS1 Antigen, IgG Antibody, IgM Antibody | Whole blood, serum, plasma | 20 | 125 |
| MP Diagnostics | MULTISURE Dengue Ab/Ag Rapid Test | 43592-020 | MPBIO_DX052016-EN-MULTISURE-Dengue-Ab-Ag-Rapid-Test-RUO-0743592-Manual MDZ0012-ENG-1, Revision 05-2016 | NS1 Antigen, IgG Antibody, IgM Antibody, IgA Antibody | Whole blood, serum, plasma | 20-25 | 25 |

Supplemental table 2. Sensitivity by serotype of dengue rapid diagnostic tests

|  | % sensitivity (95% Confidence interval) | | | |
| --- | --- | --- | --- | --- |
| **Manufacturer** | **DENV1** | **DENV2** | **DENV3** | **DENV4** |
| Abbexa | 61% (43-76) | 33% (10-65) | 68% (48-84) | 20% (4-48) |
| Biopanda | 63% (46-78) | 42% (15-72) | 61% (41-78) | 53% (27-79) |
| InBios | 68% (51-82) | 50% (21-79) | 68% (48-84) | 20% (4-48) |
| Artron | 84% (69-94) | 83% (52-98) | 89% (72-98) | 53% (27-79) |
| Cortez Diagnostics | 74% (57-87) | 42% (15-72) | 64% (44-81) | 33% (12-62) |
| Creative Diagnostics | 61% (43-76) | 25% (5-57) | 61% (41-78) | 13% (2-40) |
| CTK Biotech | 89% (75-97) | 83% (52-98) | 79% (59-92) | 47% (21-73) |
| LumiQuick Diagnostics | 82% (66-92) | 33% (10-65) | 71% (51-87) | 33% (12-62) |
| MP Diagnostics | 58% (41-74) | 0% (0-26) | 39% (22-59) | 0% (0-22) |

Supplemental Table 3. STARD Checklist

| **Section & Topic** | **No** | **Item** | **Reported on page #** |
| --- | --- | --- | --- |
| TITLE OR ABSTRACT |  |  |  |
|  | 1 | Identification as a study of diagnostic accuracy using at least one measure of accuracy  (such as sensitivity, specificity, predictive values, or AUC) | 1 |
| ABSTRACT |  |  |  |
|  | 2 | Structured summary of study design, methods, results, and conclusions  (for specific guidance, see STARD for Abstracts) | 3 |
| INTRODUCTION |  |  |  |
|  | 3 | Scientific and clinical background, including the intended use and clinical role of the index test | 4-5 |
|  | 4 | Study objectives and hypotheses | 5 |
| METHODS |  |  |  |
| *Study design* | 5 | Whether data collection was planned before the index test and reference standard  were performed (prospective study) or after (retrospective study) | 5 |
| *Participants* | 6 | Eligibility criteria | 5 |
|  | 7 | On what basis potentially eligible participants were identified  (such as symptoms, results from previous tests, inclusion in registry) | 5 |
|  | 8 | Where and when potentially eligible participants were identified (setting, location and dates) | 5 |
|  | 9 | Whether participants formed a consecutive, random or convenience series | 5 |
| *Test methods* | 10a | Index test, in sufficient detail to allow replication | 6-7 |
|  | 10b | Reference standard, in sufficient detail to allow replication | 5-6 |
|  | 11 | Rationale for choosing the reference standard (if alternatives exist) | 5 |
|  | 12a | Definition of and rationale for test positivity cut-offs or result categories  of the index test, distinguishing pre-specified from exploratory | 7 |
|  | 12b | Definition of and rationale for test positivity cut-offs or result categories  of the reference standard, distinguishing pre-specified from exploratory | 7 |
|  | 13a | Whether clinical information and reference standard results were available  to the performers/readers of the index test | 7 |
|  | 13b | Whether clinical information and index test results were available  to the assessors of the reference standard | 7 |
| *Analysis* | 14 | Methods for estimating or comparing measures of diagnostic accuracy | 7 |
|  | 15 | How indeterminate index test or reference standard results were handled | 7 |
|  | 16 | How missing data on the index test and reference standard were handled | 7 |
|  | 17 | Any analyses of variability in diagnostic accuracy, distinguishing pre-specified from exploratory | 7 |
|  | 18 | Intended sample size and how it was determined | 5 |
| RESULTS |  |  |  |
| *Participants* | 19 | Flow of participants, using a diagram | 9, Sup fig 3-11 |
|  | 20 | Baseline demographic and clinical characteristics of participants | 8-9 |
|  | 21a | Distribution of severity of disease in those with the target condition |  |
|  | 21b | Distribution of alternative diagnoses in those without the target condition |  |
|  | 22 | Time interval and any clinical interventions between index test and reference standard | 9, Sup fig 3-4 |
| *Test results* | 23 | Cross tabulation of the index test results (or their distribution)  by the results of the reference standard | 9-13, Fig 1-4 |
|  | 24 | Estimates of diagnostic accuracy and their precision (such as 95% confidence intervals) |  |
|  | 25 | Any adverse events from performing the index test or the reference standard |  |
| DISCUSSION |  |  |  |
|  | 26 | Study limitations, including sources of potential bias, statistical uncertainty, and generalisability | 14 |
|  | 27 | Implications for practice, including the intended use and clinical role of the index test | 12-14 |
| OTHER INFORMATION |  |  |  |
|  | 28 | Registration number and name of registry |  |
|  | 29 | Where the full study protocol can be accessed |  |
|  | 30 | Sources of funding and other support; role of funders | 16 |

**Supplemental Figures**

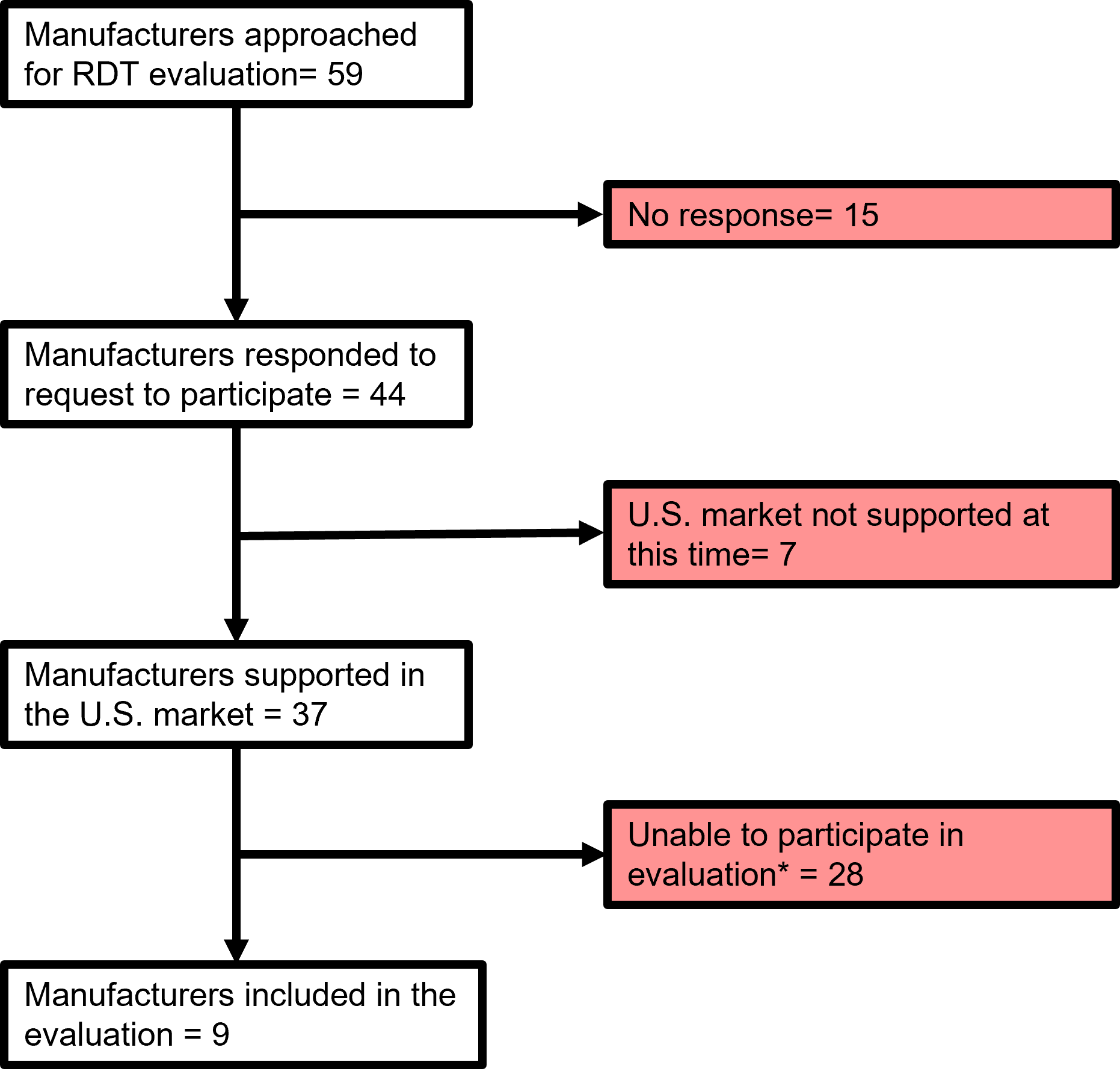

Supplemental Figure 1. Flowchart of the selection process for dengue rapid diagnostic tests (RDTs) evaluated in this study. *Manufacturers were not able to participate for a variety of reasons including requiring large minimum orders, requiring purchases through wire or bank transfer, would only sell to distributors and retailers, and no interest in participating in an evaluation.

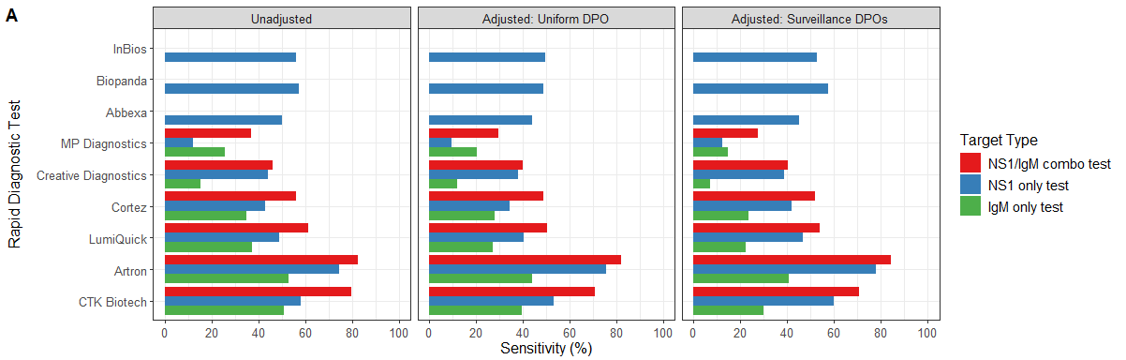

Supplemental Figure 2. Comparison of crude and adjusted sensitivity and specificity of dengue rapid diagnostic tests (RDTs), stratified by target analyte, using composite reference standard by day post-symptom onset (DPO). The composite reference standard defined dengue-positive samples as those positive by any FDA-approved RT-PCR, NS1, or IgM ELISA dengue test. Crude sensitivity and two adjusted estimates: one assuming a uniform number of tests each day post-onset (DPO) and one based on observed testing patterns in a hospital-based enhanced surveillance system in Puerto Rico. Red bars indicate NS1/IgM combination RDTs; blue bars indicate NS1-only RDTs; green bars indicate IgM-only RDTs.

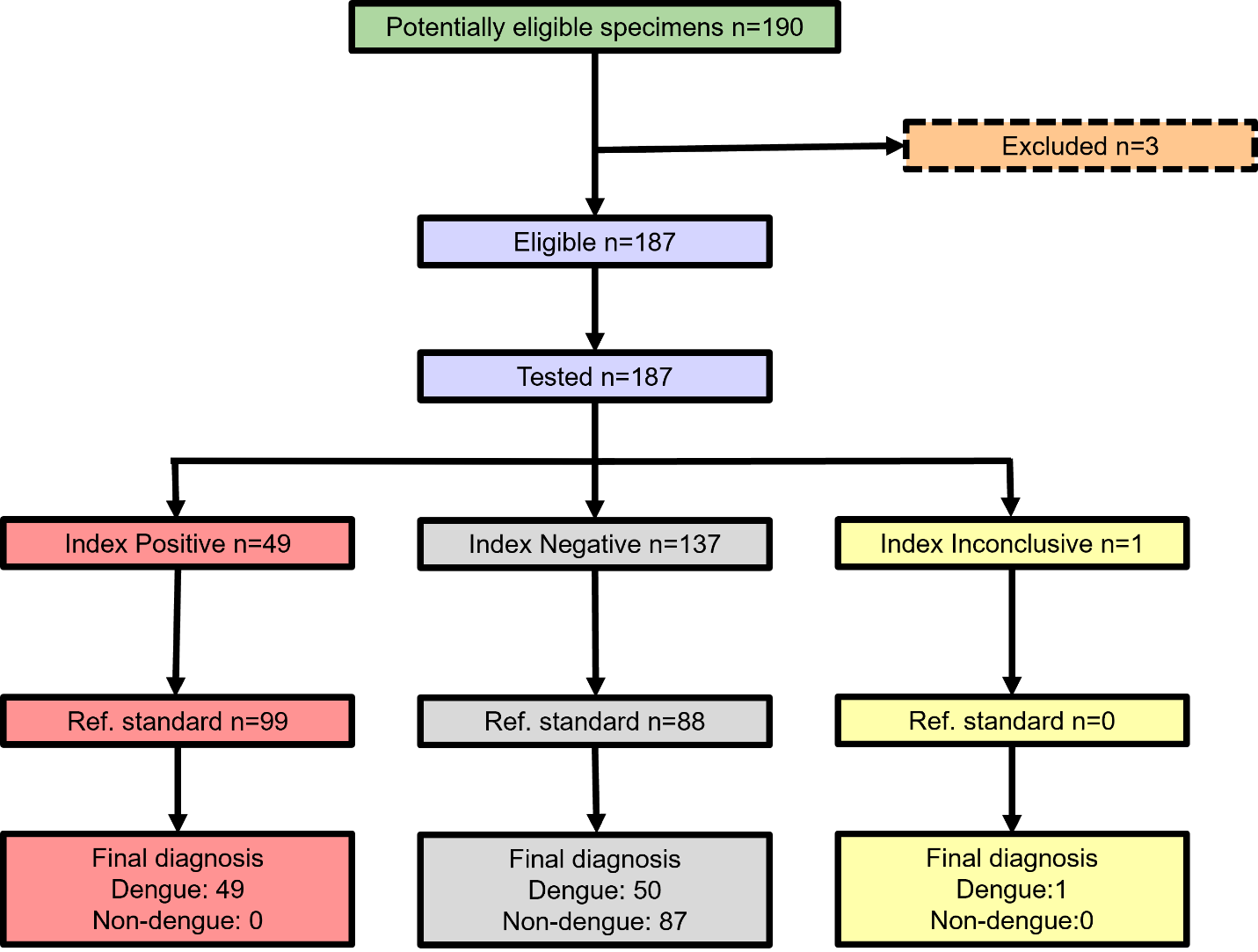

Supplemental Figure 3. Sample flow diagram for the evaluation of the Abbexa Dengue Virus NS1 Antigen Rapid Test. The index test refers to the Abbexa RDT, with final diagnostic classification based on a composite reference standard that includes rRT-PCR, NS1 ELISA, and IgM ELISA.

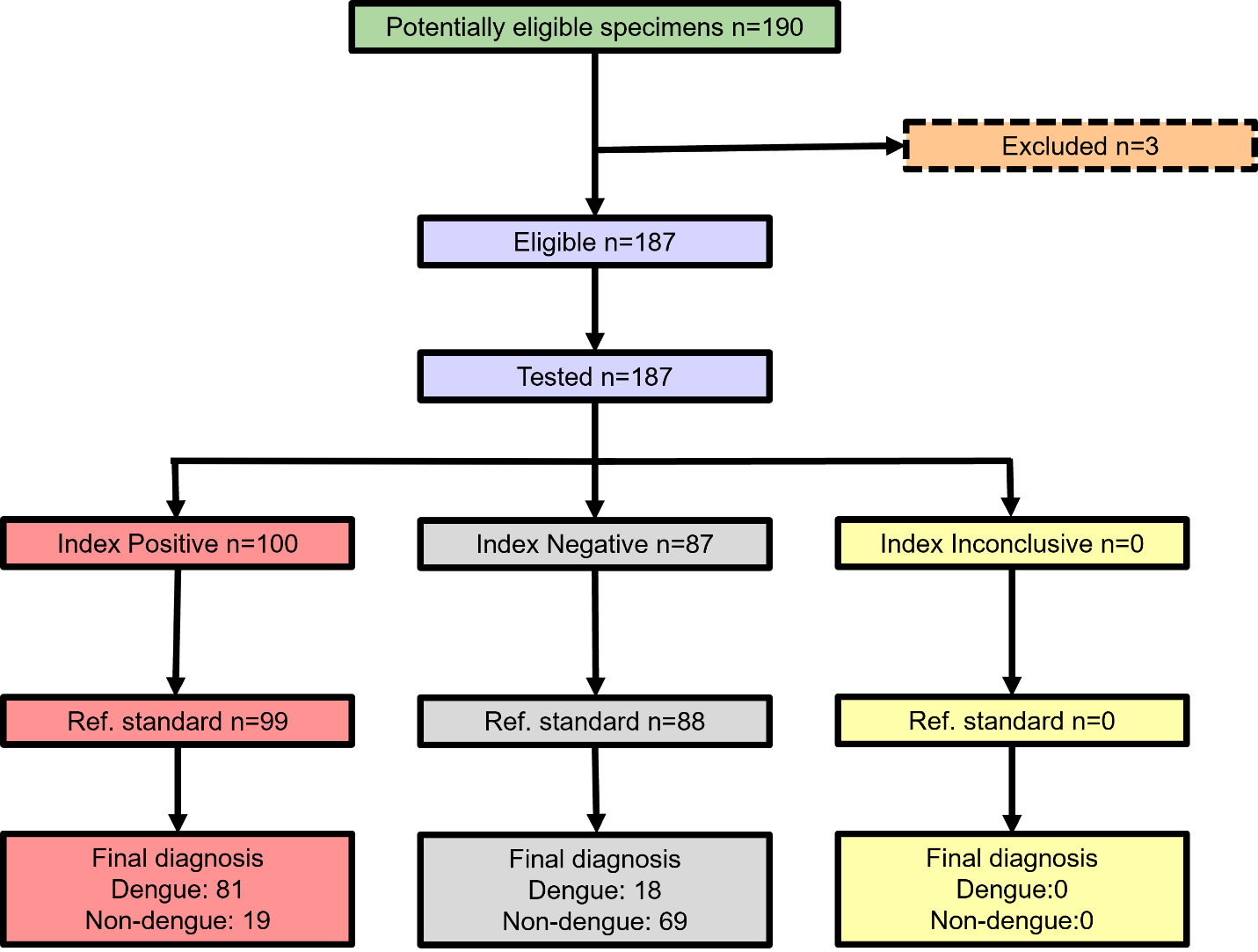

Supplemental Figure 4. Sample flow diagram for the evaluation of the Artron Dengue IgG/IgM & NS1 Test. The index test refers to the Artron RDT, with final diagnostic classification based on a composite reference standard that includes rRT-PCR, NS1 ELISA, and IgM ELISA.

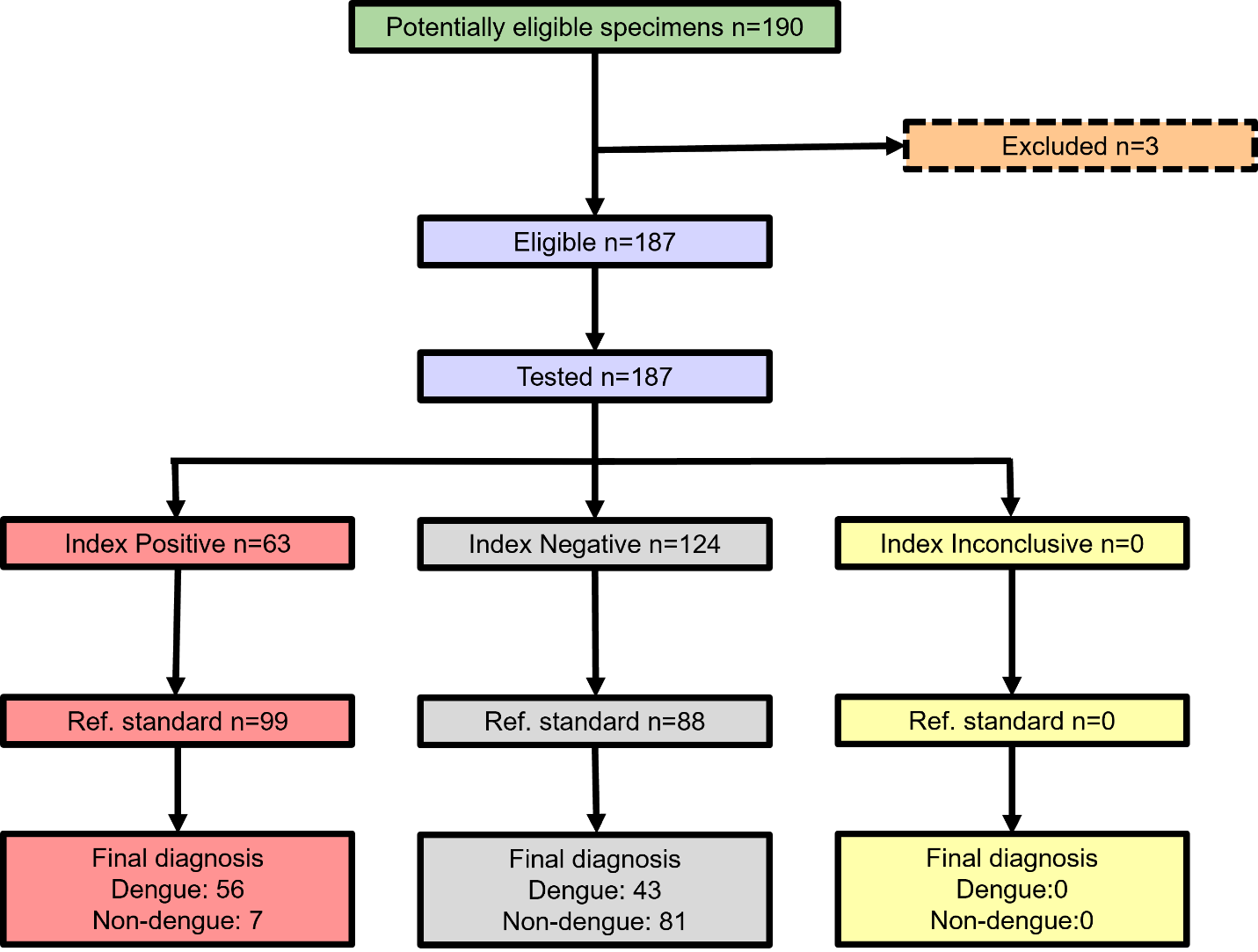

Supplemental Figure 5. Sample flow diagram for the evaluation of the Biopanda Dengue NS1 Rapid Test Cassette Test. The index test refers to the Biopanda RDT, with final diagnostic classification based on a composite reference standard that includes rRT-PCR, NS1 ELISA, and IgM ELISA.

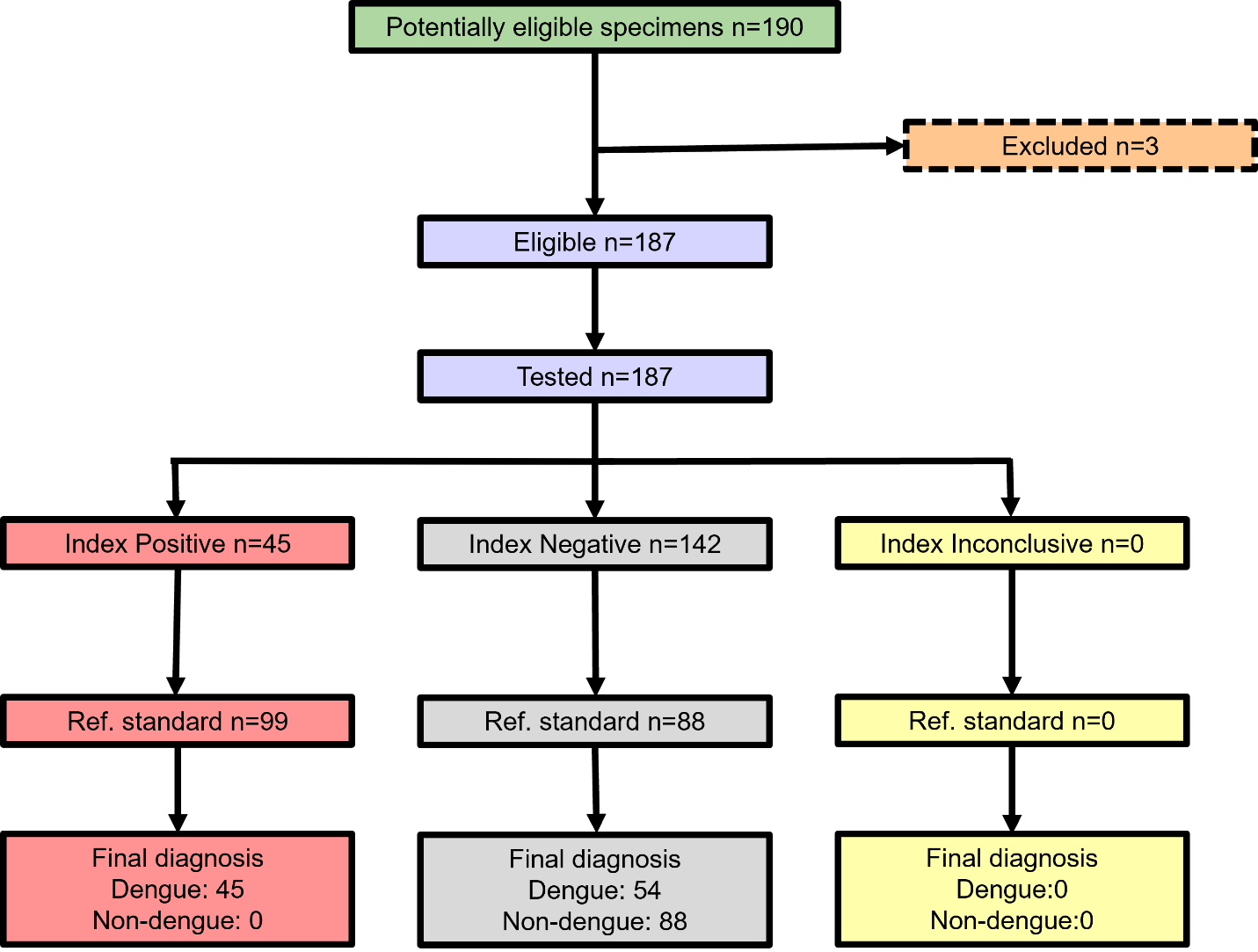

Supplemental Figure 6. Sample flow diagram for the evaluation of the Creative Diagnostics Dengue IgG/IgM/NS1 Combo Rapid Test. The index test refers to the Creative Diagnostics RDT, with final diagnostic classification based on a composite reference standard that includes rRT-PCR, NS1 ELISA, and IgM ELISA.

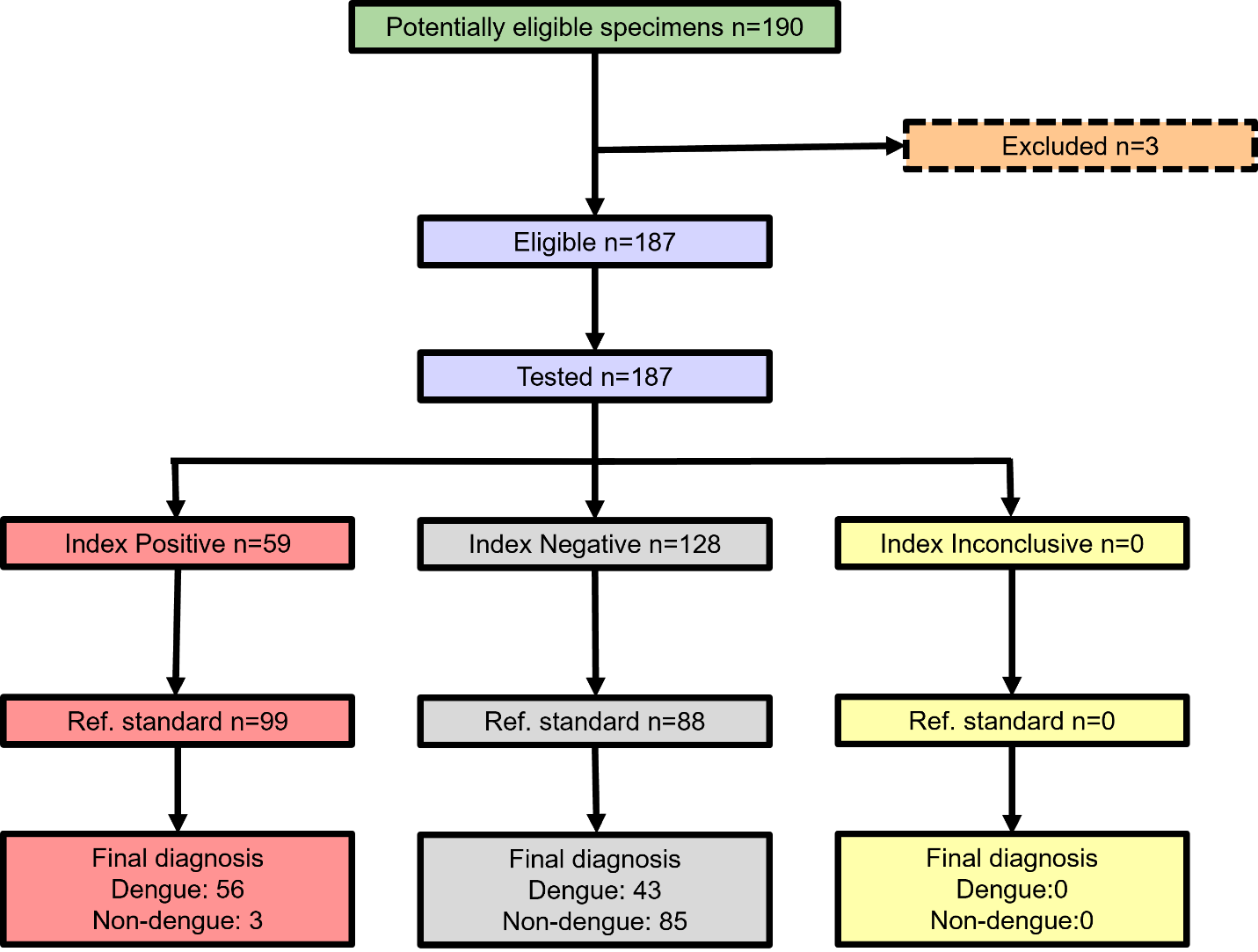

Supplemental Figure 7. Sample flow diagram for the evaluation of the Cortez Diagnostics OneStep Dengue NS1 Antigen & IgG/IgM Antibody Duo Panel Rapicard™ InstaTest. The index test refers to the Cortez Diagnostics RDT, with final diagnostic classification based on a composite reference standard that includes rRT-PCR, NS1 ELISA, and IgM ELISA.

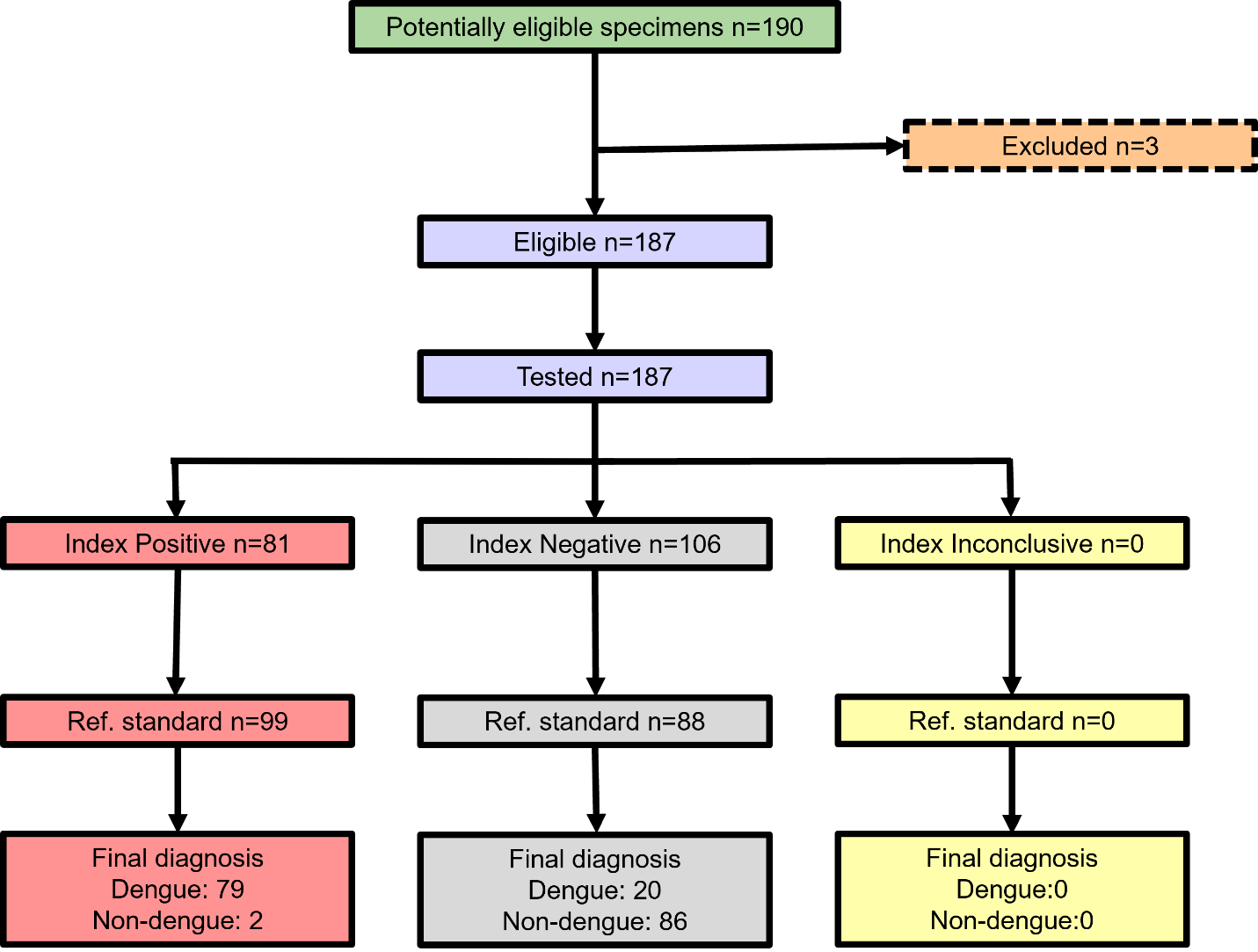

Supplemental Figure 8. Sample flow diagram for the evaluation of the CTK Biotech OnSite Duo Dengue Ag-IgG/IgM Rapid Test-Cassette. The index test refers to the CTK Biotech RDT, with final diagnostic classification based on a composite reference standard that includes rRT-PCR, NS1 ELISA, and IgM ELISA.

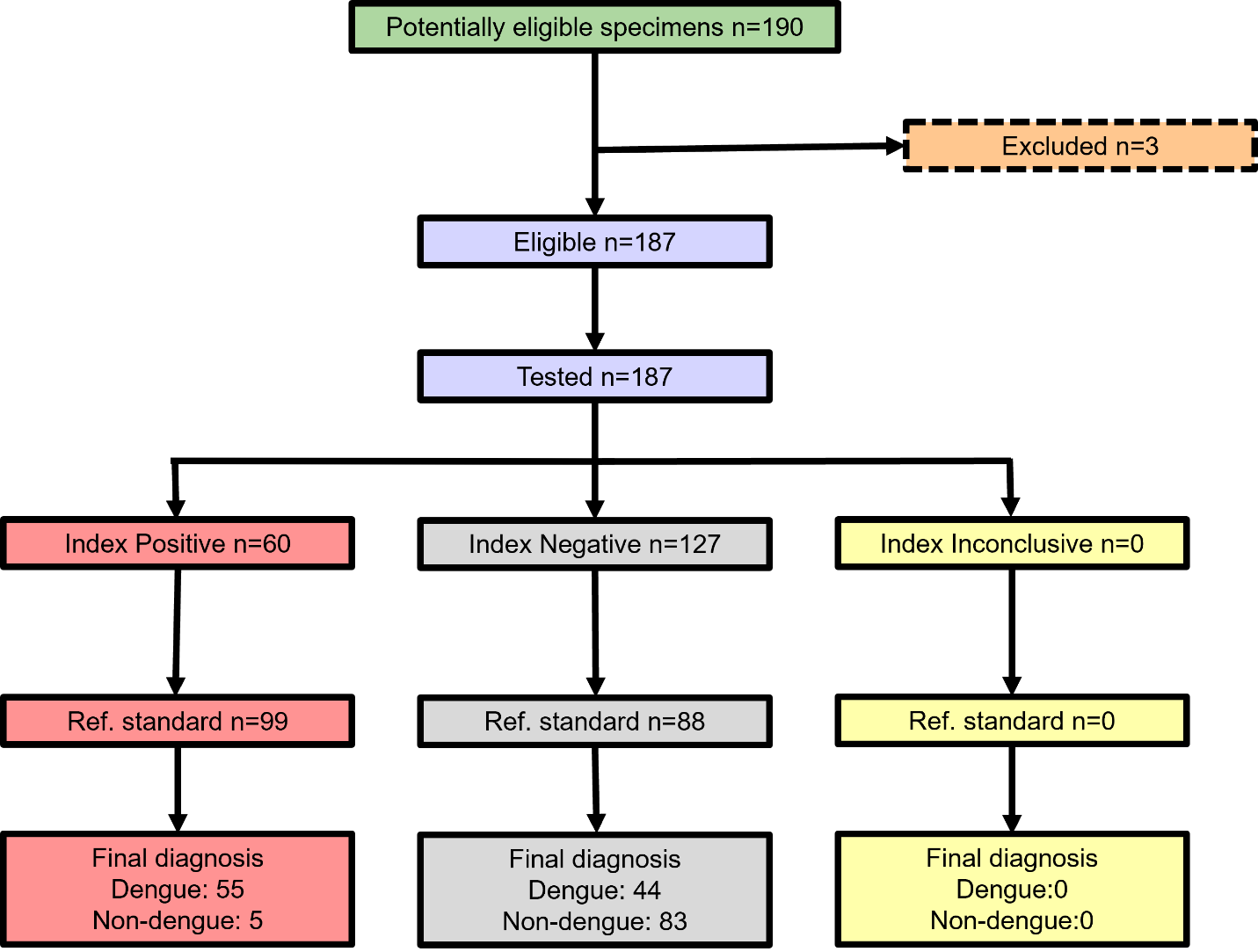

Supplemental Figure 9. Sample flow diagram for the evaluation of the InBios Dengue NS1 Detect™ Rapid Test. The index test refers to the InBios RDT, with final diagnostic classification based on a composite reference standard that includes rRT-PCR, NS1 ELISA, and IgM ELISA.

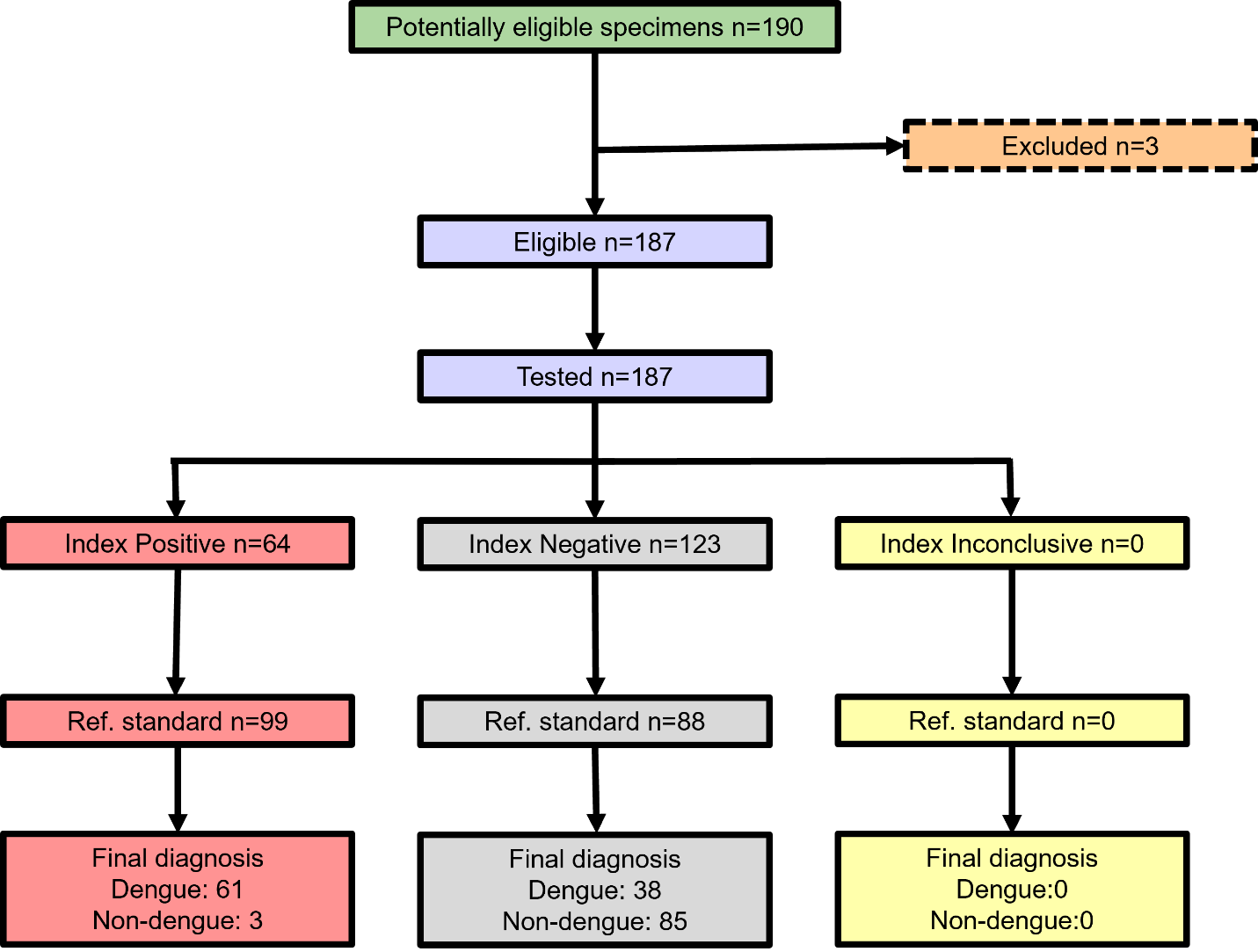

Supplemental Figure 10. Sample flow diagram for the evaluation of the LumiQuick Diagnostics Dengue NS1 Antigen & IgG/IgM Antibody Duo Panel. The index test refers to the LumiQuick RDT, with final diagnostic classification based on a composite reference standard that includes rRT-PCR, NS1 ELISA, and IgM ELISA.

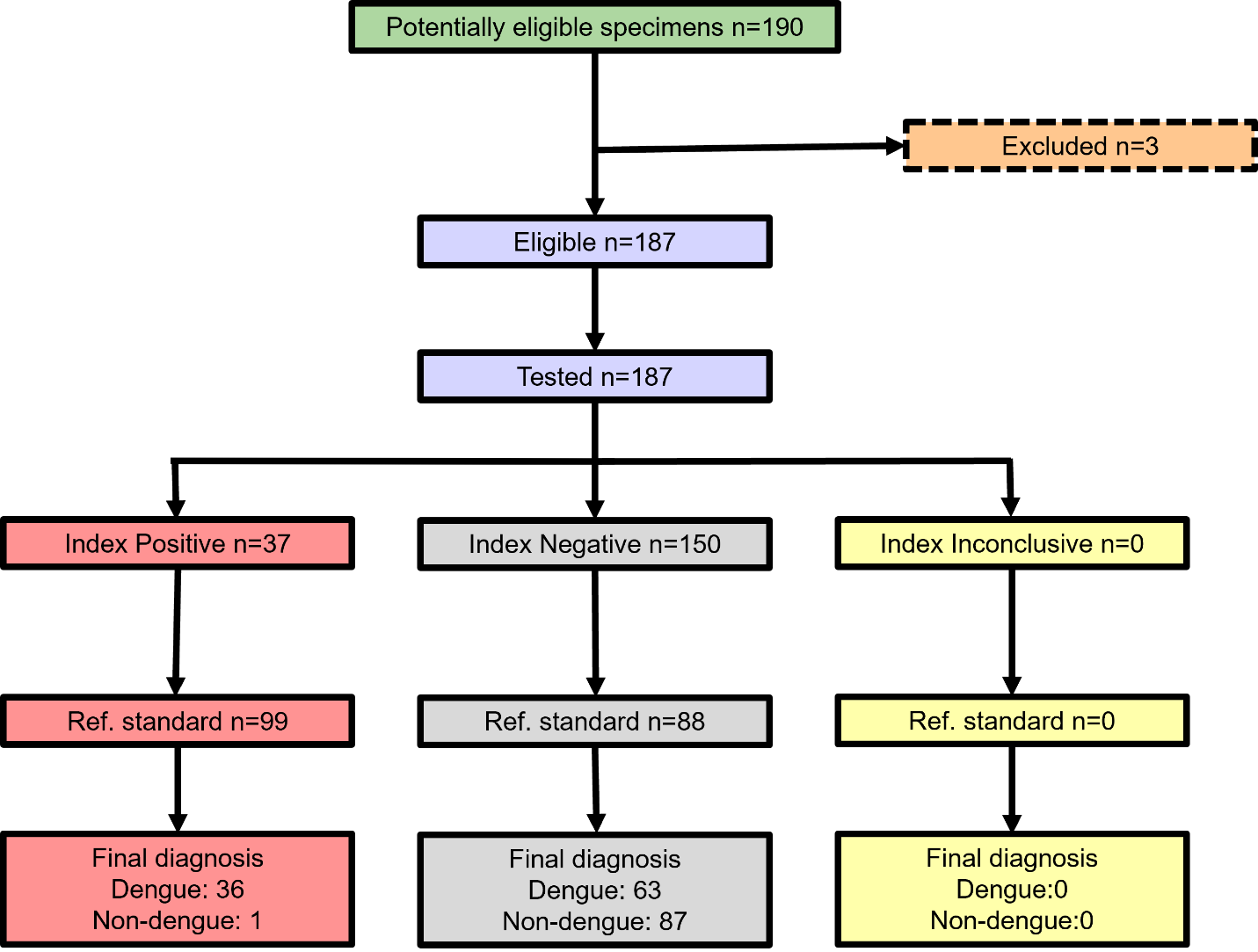

Supplemental Figure 11. Sample flow diagram for the evaluation of the MP Diagnostics MULTISURE Dengue Ab/Ag Rapid Test. The index test refers to the MP RDT, with final diagnostic classification based on a composite reference standard that includes rRT-PCR, NS1 ELISA, and IgM ELISA.
